# Effectiveness of Leading Pedestrian Intervals for Pedestrian Safety in New York City

**DOI:** 10.1101/2024.05.03.24306847

**Authors:** Siddhesh Zadey, Leah E. Roberts, Brady Bushover, Ariana N. Gobaud, Christina A. Mehranbod, Carolyn S. Fish, Evan L. Eschliman, Xiang Gao, Dana E. Goin, Christopher N. Morrison

## Abstract

Road traffic crashes are a leading cause of morbidity and mortality in large cities. Cities including New York City (NYC) in the United States (US) have implemented a suite of interventions to reduce road traffic crashes. Leading pedestrian intervals (LPIs) are a low-cost physical environmental intervention premised to reduce vehicle-pedestrian crashes by providing pedestrians a head-start over the turning vehicular traffic. Using a spatial ecological panel design, we assessed the impact of LPIs on the risk of total, non-fatal, and fatal pedestrian injuries in NYC from January 2013 to December 2018. For the 36,102 intersection-years studied and 6,017 intersections (2,883 LPI-treated) included in the study, there were 26,033 total injuries of which 291 were fatal. Significant reductions were observed for total (19.4% [95% CI: 11.8%, 26.4%]) and non-fatal injuries (19.6% [11.9%, 26.5%]) but not for fatal injuries. Multiple sensitivity analyses ensured robustness of findings. LPIs may prove to be an effective intervention to improve pedestrian safety outcomes across cities.

## Main

In 2019, road traffic crashes lead to over 1.35 million deaths and 50 million injuries globally^1^. Of these, about 23% of injuries and 37% of deaths involve pedestrians^2^. In the United States (US), over 68,000 pedestrians died and 6.10 million pedestrians were severely injured during 2011-2020^3^. Large cities like New York City (NYC) bear the majority of this injury and mortality burden, in part due to their high population densities, greater traffic flows, and more regions with high pedestrian density^4^.

NYC was among the first adopters of the US Vision Zero program — a series of preventive interventions to reduce road traffic crash-related morbidity and mortality^5^. Since 2014, NYC has spent over $850 million on preventive interventions, including changes to the physical environment^6^. Physical environmental interventions are structural changes to roadways and surroundings intended to reduce pre- and peri-event risk of an injury. Examples of such interventions include speed humps, red light cameras, turn traffic calming, and enhanced crossing, among others. Particularly relevant interventions for crashes involving pedestrians are leading pedestrian intervals (LPIs) that are traffic light sequences providing pedestrians with a brief head start (7-11 seconds for NYC) to begin crossing an intersection before the turning vehicular traffic is permitted to enter. Since LPIs are located at intersections, their preventive mechanism is directly tied to crashes at and around intersections.

LPIs are one of the most low-cost and easily implementable interventions in Vision Zero. There is mixed evidence on the association of LPIs in twelve US states with reduced crashes and injuries^7^. Studies on association between LPI and reduced pedestrian crashes and LPIs have relied on small number of intersections and short observation windows of time, reducing study power, reliability, and generalizability^8–10^. Further, the commonly used study designs lack appropriate comparison groups since untreated intersections may be systematically different than treated intersections by various factors that affect LPI implementation priority. Studies have also missed investigating the effects at a high spatial resolution that accords with the theoretical causal mechanism (i.e., the intersection level). Hence, a rigorous evaluation of LPI effectiveness is missing.

Our study employs a rigorous spatial ecological panel design to test the effectiveness of leading pedestrian intervals in NYC in reducing the risk of fatal and non-fatal pedestrian injuries at and around intersections. With 6017 intersections, NYC provides the largest possible dataset to estimate effectiveness of LPIs. Our study uses study design, spatial scale, and statistical methods that can simultaneously account for multiple threats to internal validity.

## Results

From January 2013 to December 2018, about 48% of the 6,017 intersections included here had LPIs installed. **Supplementary Table 1** provides the distributions of outcomes across treated and control units (i.e., intersection-years). Nineteen percent of the 36,102 intersection-years were treated units. For both treated and control units, 341,882 pedestrian-involving crashes at and around intersections led to 26,033 non-fatal and fatal pedestrian injuries (1.1% fatalities). LPI-treated intersection-years saw 20.3% of fatal and 18.7% of non-fatal injuries **(Supplementary Figures 1a-d)**.

The primary analysis found a significant 19.4% (95% CI: 11.8%, 26.4%) reduction in the risk of total injuries at LPI-treated units compared to those without LPIs **(Figure 1)**. With a baseline risk of 42.2% among the control units, the corresponding absolute risk reduction was 4 total injuries per 100 intersection-years **(Supplementary Table 2)**. A significant reduction of 19.6% (11.9%, 26.5%) was also observed for non-fatal injuries. With a baseline risk of 41.9%, the corresponding absolute risk reduction was 4 non-fatal injuries per 100 intersection-years. Sensitivity analyses confirmed these findings for total and non-fatal injury risk. LPIs did not significantly reduce the risk of fatal injuries **(Figure 1)**. Further, the point estimate for the fatal injury risk was unstable across primary and sensitivity analyses (aOR=1.1 vs. 0.7, respectively).

**Figure 1:**
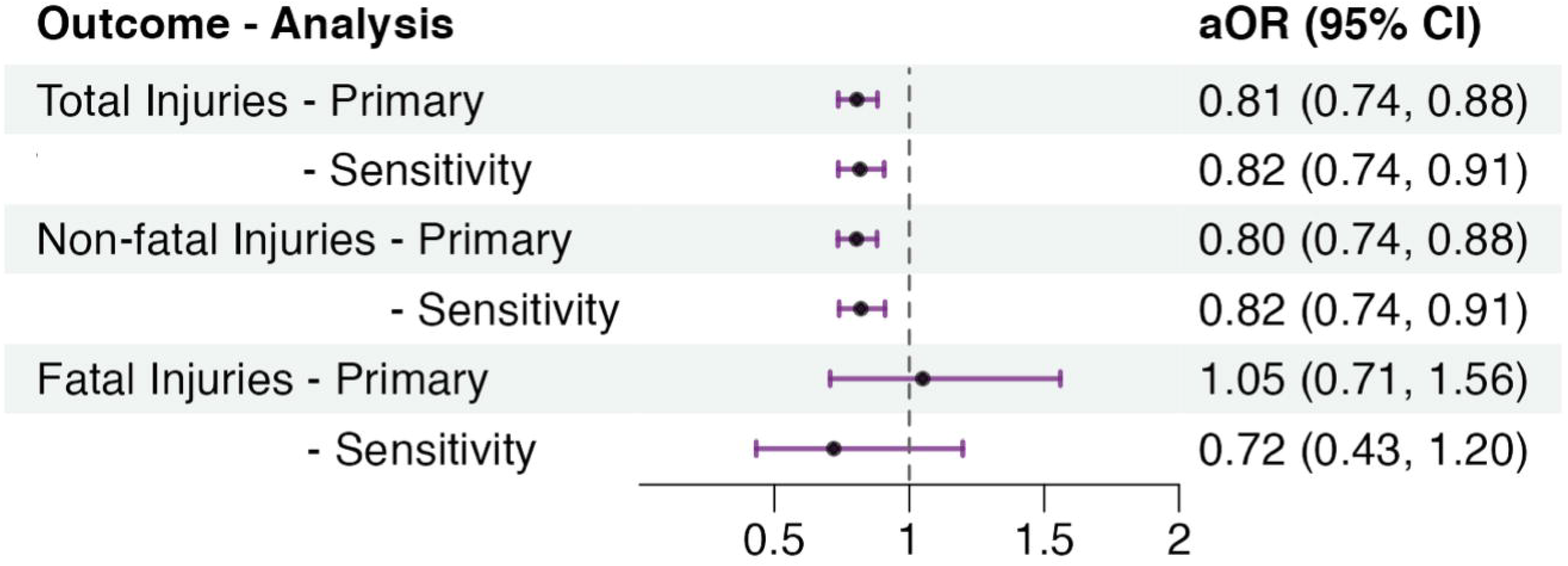
Forest plot for effectiveness (adjusted odds ratio or aOR values) of LPI on total, non-fatal, and fatal pedestrian injuries. The models have fixed effects for years and random effects for intersections.

After testing variables hypothesized to produce different strengths of effect, we did not find significant effect measure modification by the number of roadway segments crossing the intersection location, sum of segments widths, or the maximum length of the segments for the risk of total pedestrian injuries **(Table 1)**.

**Table 1:** Effect measure modification with adjusted odds ratios for the interaction terms. Models use total pedestrian injuries as the outcome and have fixed effects for years and random effects for intersections along with ‘ever treated’ and ordinal effect measure modifier terms. We used ‘crossing the intersection location’ since the spatial join cannot discriminate between small number of roadway segments including bridges or tunnels that might pass over or under the location without physically joining the intersection.

| **Effect Measure Modifier** | **aOR (95% CI)**<br>**for interaction**<br>**term** |
| --- | --- |
| No. of roadway segments crossing the intersection location |  |
| Ref (3 segments) |  |
| 4 segments (cross-roads) | 0.78 (0.59, 1.01) |
| >4 segments | 0.90 (0.67, 1.20) |
| Sum of roadway segment widths crossing the intersection location |  |
| Ref (Quartile 1) |  |
| Quartile 2 | 1.04 (0.83, 1.30) |
| Quartile 3 | 1.08 (0.86, 1.34) |
| Quartile 4 | 1.02 (0.83, 1.25) |
| Maximum length of roadway segments crossing the intersection location |  |
| Ref (Quartile 1) |  |
| Quartile 2 | 0.95 (0.77, 1.19) |
| Quartile 3 | 0.89 (0.73, 1.08) |
| Quartile 4 | 0.84 (0.68, 1.03) |

## Discussion

LPIs appear to reduce total pedestrian injuries at and around intersections in New York City by 19.5%. The sensitivity and effect measure modification analyses confirmed findings’ robustness and generalizability across intersections in NYC. Hence, LPIs are effective in NYC.

While the LPIs were more effective at intersections with crossroads than those with three-ways, this difference was not statistically significant. There might be a potential dose-response relationship with a greater effectiveness of LPIs at intersections adjacent to the longer roadway segments. However, the overlapping confidence intervals across quartiles of maximum roadway segment lengths show that such relationship is not statistically significant. The large sample ensured enough power such that absence of any statistically significant interactions depict an absence of effect measure modification. Hence, LPIs are effective regardless of street-level characteristics of the roadway segments.

Results of this study accord with findings from prior evaluations on association of LPIs with reduced pedestrian crash risk. Previous estimates for LPIs have varied from 12% to 95% reduction in pedestrian-involved crashes across different cities, outcomes measures, and analytical methods^7^. However, the majority of studies relied on a small number of intersections (n=3–105). Specifically, for NYC, three past studies conducted at different times from the mid-1990s to mid-2010s noted 28% to 37% reductions using 26–104 intersections^8–10^. In contrast, we used the largest yet sample of intersections (n=6,017), allowing us to produce more precise effect size estimates. Previous studies used pre-post or case-control designs^8–10^. Our rigorous design ensured better comparability between treated and control units and the model specification allowed for random effects across intersections. Further, using intersection as the geographic unit of analysis is line in with the mechanism of action of LPIs.

The current study has several limitations. First, the temporal unit of analysis (years) may have led to misclassification of outcome occurrence relative to time of treatment. However, the year-lagged sensitivity analysis conducted showed robustness of effect for total and non-fatal injuries. Further, selecting years helped obviate seasonality and temporal autocorrelation in outcomes. Second, spatial autocorrelation may have biased the estimates. However, we used the spatial buffers to include only the crashes within 100 feet of the intersections, which ensured independence of intersections from one another. Third, unmeasured confounding is plausible. However, the estimated effect that was large enough, which is unlikely to be impacted by such confounding. Fourth, our estimate for fatal injuries was neither stable across primary and sensitivity analyses nor significant. This could depict reverse causation as LPI interventions are installed at intersections that observed a fatal injury in the previous year. Future studies using pooled data across cities could provide more reliable estimates for sparse outcomes such as fatal injuries. Finally, the findings may not be transportable to other settings including smaller cities, given the uniqueness of the NYC context. However, we estimated effects at the intersection level and also assessed that these are consistent with the assessed effect measure modifiers, marking the generalizability to other parts of NYC and cities with similar pedestrian and vehicular traffic volumes.

## Conclusion

We present a novel intersection-level assessment of New York City demonstrating that leading pedestrian intervals are effective in reducing the risk of pedestrian injuries at and around intersections for reducing non-fatal injuries. Our analysis supports implementing LPIs at about 30,000 currently untreated intersections in NYC. LPIs might also be useful in other large cities with high traffic volumes with increased risk for pedestrian-involving crashes at intersections. Future work should test whether bundling LPIs with other physical environmental interventions, such as turn traffic calming, enhanced crossing, etc. at and around intersections has synergistic protective effects.

## Methods

The study setting was NYC from January 2013 to December 2018. We used a spatial ecological panel design to assess the risk reduction in pedestrian injuries, with intersection-year as the unit of analysis (see **Supplementary Methods** for details). NYC Open Data and NYC Vision Zero websites provided precise geolocations (latitude and longitude coordinates) of outcome and exposure/intervention data^11^.

### Variables

#### Outcomes

We considered non-fatal and fatal pedestrian injuries in collisions with motor vehicles within 100 feet of an intersection in a year as outcomes. We conducted spatial joins between crash and intersection files to filter the eligible crashes. We chose a 100-feet buffer as it approximates the size of an intersection and it is likely causally associated with the intersection traffic flow.

#### Treatment

We considered roadway intersections with LPIs within 10 feet as treated, while those without an LPI within 10 feet were considered untreated. Installment year denoted treatment initiation. Hence, the treatment status was dependent on both location and time.

We included intersections that had LPIs installed during January 2013 – December 2023. We defined intersections where LPI installation occurred from January 2013 – December 2018 as treatment sites. We designated those with installations occurring after December 2018 as control sites. Our approach provided comparability between treatment and control (not yet treated) sites insofar as NYC noted them as eligible and needing LPIs.

#### Effect Measure Modifiers

We considered three modifiers: number of roadway segments crossing the intersection location (ordinal coding of 3, 4, and >4 segments), sum of segments widths (coded as quartiles), and maximum length of the segments (coded as quartiles).

### Analysis

We used mixed effects logistic regression to estimate the treatment effect of LPIs in separate models for the odds of total, non-fatal, and fatal injuries. Along with the treatment variable of interest, all models included fixed effects for years, random effects (intercepts) for intersections, and a term for the ‘ever treated’ status of the intersection.

The adjusted odds ratios (aORs) approximate the relative risk since outcome count at each intersection was relatively rare. Robust standard errors account for heteroskedasticity across intersection sites. We estimated absolute risk reduction (ARR) per intersection-year using the aORs and baseline risk estimates in the control group.

We conducted a sensitivity analysis to test for temporal misspecification of the outcome events relative to LPI installment using a year-lagged treatment variable. For example, for an intersection with LPI installed in 2014, we considered the treatment effect for 2015 and beyond.

We conducted effect measure modification analysis using similar mixed effect models for total injury risk as the outcome and interpreted the aORs of the interaction terms to investigate significant effect modification. The ordinal coding ensured model parsimony. Use of quartiles ensured that the outliers in the distribution of a modifier did not skew the effect.

We used ArcGIS Pro 3.0.0 for geospatial data preprocessing, Stata 18.0 (College Station, Texas) for statistical modeling, Datawrapper for visualization, R 4.0.2 for data wrangling, modeling, and visualization, and Microsoft® Excel 16.29 for validation.

## Supporting information

Supplement main file

## Data Availability

All data produced in the present study are available upon reasonable request to the authors

## Acknowledgments

We thank Dr. Lauren C. Houghton, Mr. Alex Furuya, Mr. Marco Thimm-Kaiser, Ms. Michelle L. Lui, and Ms. Navjot Buttar for their helpful feedback.

## Ethics Statement

We used publicly available event-level data with no personally identifiable information. Hence, the study did not need an IRB review or approval.

## Data Sharing Statement

Data used in this study is publicly available at NYC Open Data while that generated can be requested from the authors. NYC Open Data (https://data.cityofnewyork.us/Public-Safety/Motor-Vehicle-Collisions-Crashes/h9gi-nx95) provided day-by-day incident-level data for all motor vehicle crashes since 2012. The NYC Vision Zero website provided data on LPI locations and installation dates (https://data.cityofnewyork.us/Transportation/VZV_Leading-Pedestrian-Interval-Signals/mqt5-ctec).

