## Supplement main file for "Effectiveness of Leading Pedestrian Intervals for Pedestrian Safety in New York City"

**Supplementary Figure**

**[Include figure here]**

**Supplementary Figure 1a-d: a) Street view map of New York City showing geolocated intersections with and without leading pedestrian intervals from January 2013 to December 2018. Crashes at and around intersections (within 100 feet) involving pedestrians that led to b) total, c) non-fatal, and d) fatal injuries.** Blue and red dots represent treated and control (not yet treated) intersections, respectively—distance buffer for linking a leading pedestrian interval installation to an intersection: 10 feet. Bubbles size depicts outcome count at the intersection.

**Supplementary Tables**

**Supplementary Table 1: Study units and outcomes by treatment and control groups.**

|  | **Total** | **LPI-Treated** | **Control** |
| --- | --- | --- | --- |
| Intersections | 6,017 | 2,883 | 3,134 |
| Units | 36,102 | 6,685 | 29,417 |
| Pedestrian involving crashes | 341,882 | 6,6119 | 275,763 |
| Total Pedestrian Injuries | 26,033 | 4,890 | 21,143 |
| Non-fatal Injuries | 25,742 | 4,831 | 20,911 |
| Fatal injuries | 291 | 59 | 232 |

**Supplementary Table 2: Absolute risk reduction and baseline risks across treatment and control groups (shown per intersection-year).**

| **Outcome - Analysis** | **ARR (95% CI)** | **Base risk in treatment group (95% CI)** | **Base risk in control group (95% CI)** |
| --- | --- | --- | --- |
| Total Injuries - Primary | -0.0406 (-0.0573, -0.0239) | 0.3810 (0.3657, 0.3963) | 0.4216 (0.4137, 0.4296) |
| Total Injuries - Sensitivity | -0.0258 (-0.0448, -0.0067) | 0.3908 (0.3723, 0.4093) | 0.4166 (0.4090, 0.4241) |
| Non-fatal Injuries - Primary | -0.0409 (-0.0575, -0.0242) | 0.3778 (0.3626, 0.3931) | 0.4187 (0.4108, 0.4266) |
| Non-fatal Injuries - Sensitivity | -0.0252 (-0.0441, -0.0062) | 0.3884 (0.3700, 0.4068) | 0.4135 (0.4060, 0.4211) |
| Fatal Injuries - Primary | 0.0004 (-0.0028, 0.0035) | 0.0082 (0.0055, 0.0110) | 0.0078 (0.0067, 0.0089) |
| Fatal Injuries - Sensitivity | -0.0010 (-0.0044, 0.0024) | 0.0070 (0.0038, 0.0102) | 0.0080 (0.0070, 0.0090) |

**Supplement Text**

**Supplementary Methods**

***Study Design Rationale***

Our quasi-experimental technique relied on more lenient exchangeability criteria to infer causal effects from observational longitudinal data. Using intersections provided a high geographic resolution that is in line with the mechanism of action. Using year avoided seasonality and was suitable in this case given the sparse outcome data across treated and control intersections; this would be applicable especially for fatal injuries.

**Supplementary Results**

**Supplementary Figures 1a** shows the geospatial distribution of LPI-treated (blue) and control units (red) across NYC. While **Supplementary Figures 1b-d** depict the geographic distribution of total, non-fatal, and fatal injuries over the study period across LPI-treated and control intersections. Larger bubbles depict greater counts of injuries with sparse data for fatal injuries.
