## Supplementary figures and images for "Effectiveness of Leading Pedestrian Intervals for Pedestrian Safety in New York City"

### Supplement figure

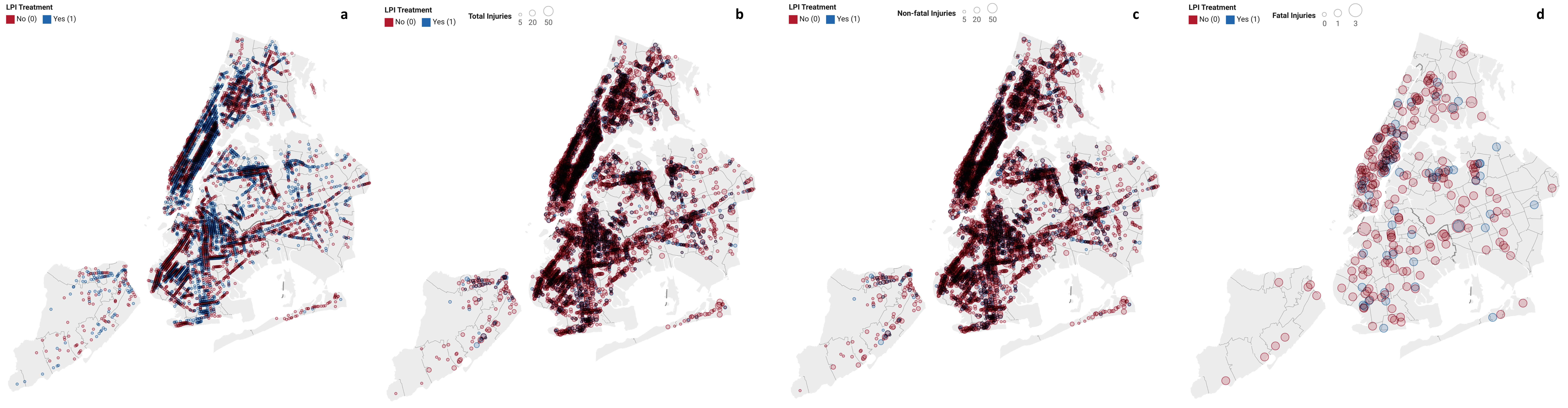
